# Biallelic Variants in *CCN2* Underlie an Autosomal Recessive Kyphomelic Dysplasia

**DOI:** 10.1101/2024.04.05.24305189

**Authors:** Swati Singh, Sumita Danda, Neetu Sharma, Hitesh Shah, Vrisha Madhuri, Mir Tariq Altaf, Nadia Zipporah Padala, Raghavender Medishetti, Alka Ekbote, Gandham SriLakshmi Bhavani, Aarti Sevilimedu, Katta M Girisha

## Abstract

Kyphomelic dysplasia is a rare heterogenous group of skeletal dysplasia, characterized by bowing of the limbs, severely affecting femora with distinct facial features. Despite its first description nearly four decades, the precise molecular basis of this condition remained elusive until the recent discovery of *de novo* variants in the KIF5B-related kyphomelic dysplasia. We ascertained two unrelated consanguineous families with kyphomelic dysplasia. They had six affected offsprings and we performed a detailed clinical evaluation, skeletal survey, and exome sequencing in three probands. All the probands had short stature, cleft palate, and micro-retrognathia. Radiographs revealed kyphomelic femora, bowing of long bones, radial head dislocations and spondyloepimetaphyseal dysplasia. We noted two novel homozygous variants in *CCN2* as possible candidates that segregated with the phenotype in the families: a missense variant c.443G>A; p.(Cys148Tyr) in exon 3 and a frameshift variant, c.779_786del; p.(Pro260LeufsTer7) in exon 5. *CCN2* is crucial for proliferation and differentiation of chondrocytes. Earlier studies have shown that *Ccn2*-deficient mice exhibit twisted limbs, short and kinked sterna, broad vertebrae, domed cranial vault, shorter mandibles, cleft palate and impaired osteoclastogenesis. We studied the impact of *CCN2* knockout in zebrafish models via CRISPR-Cas9 gene editing. F0 knockouts of *ccn2a* in zebrafish showed altered body curvature, impaired cartilage formation in craniofacial region and either bent or missing tails recapitulating the human phenotype. Our observations in humans and zebrafish combined with previously described skeletal phenotype of *Ccn2* knock out mice, confirm that biallelic loss of function variants in *CCN2* result in an autosomal recessive kyphomelic dysplasia.

## Introduction

Kyphomelic dysplasia represents a heterogenous group of rare genetic skeletal disorders, characterized by incurvation of limbs primarily affecting the femora, along with short stature, short and wide iliac wings, horizontal acetabular roof, platyspondyly, metaphyseal flaring and distinctive facial features that include prominent forehead, micrognathia, microstomia, cleft palate and low set ears. ^1–4^ The term “kypho” originates from ancient Greek word “kyphos” meaning “bent”, while “melia” refers to “limb”. The term ‘kyphomelia’ was first used to describe a skeletal dysplasia by MacLean in 1983,^1^ noting an infant with broad and severely angulated short femora, congenital bowing of other long bones, narrow thorax, platyspondyly, micrognathia, and skin dimples while also comparing the clinical findings in four patients reported earlier. ‘Kyphomelic dysplasia’ was used to contrast this phenotype from ‘campomelic dysplasia’ that has less acute bending of femora.

In the recent nosology of genetic skeletal disorders,^5^ kyphomelic dysplasia is categorized within the bent bone dysplasia group among other entities namely, Campomelic dysplasia (OMIM# 114290), Cumming syndrome (OMIM# 211890), and Stuve–Wiedemann syndrome (OMIM# 601559), Kyphomelic dysplasia with facial dysmorphism, KIF5B related (OMIM# 614592), Bent bone dysplasia, FGFR2 related (OMIM# 614592), Bent bone dysplasia, LAMA5 related (OMIM# 620076). Kyphomelic dysplasia can be misdiagnosed as campomelic dysplasia, where mild bowing of the femur and severe anterior bowing of the tibia are consistently observed or any other disorder listed here.^6,7^ Conventionally, it was considered an autosomal recessive condition.^2,8^ Recently, heterozygous *de novo* variants in KIF5B have been found to be associated with kyphomelic dysplasia,^4^ while several bent bone dysplasias do not have a known genetic basis.

Cellular communication network factor 2 (CCN2), also referred to as connective tissue growth factor (CTGF), facilitates interactions with growth factors, cell surface proteins and extracellular matrix components. It is a matricellular protein crucial for proper skeletal growth and development, playing a vital role in regulating the differentiation and function of osteoblasts and osteoclasts.^9,10^ In vitro studies on *CCN2* demonstrated that it promotes DNA synthesis in chondrocytes.^11^ Investigations on *Ccn2* deficient mice showed broader vertebrae, shortened and kinked sterna, as well as bending in the radius, ulna, tibia, and fibula. Additionally, they exhibit abnormalities in ethmoid, a domed cranial vault, secondary cleft palate, and shortened mandibles.^12^

We evaluated three probands from two unrelated consanguineous families with bowed long bones, particularly severe in femora, and identified biallelic disease-causing variants in *CCN2,* segregating in a recessive manner. We investigated consequences of loss of function of *CCN2* activity in vivo in zebrafish models.

## Methods

### Participants

Three individuals with kyphomelic dysplasia, from two unrelated families were recruited in the study. Comprehensive medical history, clinical assessments, and radiological findings of all affected individuals were documented. Written informed consents were obtained from the affected individuals and their family members. The study has received the approval from the institutional ethics committee of Kasturba Medical College and Hospital, Manipal.

### Genetic testing

Peripheral blood (2-5 ml) was collected from the probands, siblings and their parents. The genomic DNA was isolated using QIAmp DNA Blood Mini kit (QIAGEN, Hilden, Germany) following the standard protocols. Exome sequencing was performed for all the three affected individuals with kyphomelic dysplasia. Exome sequencing utilized the Agilent sure select TWIST V6 exome capture kit (TWIST Biosciences, South San Francisco, California, USA) and run on HiSeq2000 platform (Illumina San Diego, CA, USA), achieving an average coverage of 100X, with >95% of bases covered at a minimum of 20X and 97% sensitivity. Raw data was retrieved in FASTQ format and aligned to the GRCh38 assembly using Burrows-Wheeler Aligner (v2.2.1) and our in-house pipeline based on the ‘Genome Analysis Toolkit Best Practices’.^13^ Subsequently, the data underwent annotation by ‘Annotate Variation (ANNOVAR)’,^14^ supplemented by our in-house scripts.^15^ Filtered variants were further analyzed using *in silico* pathogenicity prediction tools (such as CADD Phred, MCAP, Mutation Taster, REVEL, SIFT Indel, AlphaMissense) to assess their potential impact. Conservation analysis using the Clustal Omega tool^16^ was performed to assess the conservation of the amino acid residue across species. The allele frequency of the identified rare variant was estimated from gnomAD (V3.1.2), and our in-house data of 3188 exomes.

Sanger sequencing was performed to validate and segregate identified candidate variants in the proband and their family members. Primers were designed using Primer3 software. The sequencing was carried out on an ABI 3500 Genetic Analyzer, according to manufacturer’s protocol. The variants were described according to Human Genome Variation Society (HGVS) nomenclature, with NCBI reference sequences (NM_001901.4, NP_001892.2). Both variants were submitted to the Leiden Open Variation Database (LOVD) database (variant ID: 0000972076; 0000972075).

### Design and synthesis of Single guide RNA (sgRNA), and microinjection

For generation of *ccn2a* F0 knockout in zebrafish using CRISPR/Cas9 mediated gene editing, target regions in exon 3 (GACTGCCCAATGCCCCGCA) and exon 4 (GGCGAGACTGCTTCTCAAGG) of *ccn2a* were chosen using Benchling software. A non-target sequence not present in the zebrafish genome was chosen as a control (NT).^17^ Single guide RNA (sgRNA) template for each, was ordered as single stranded DNA with the addition of T7 promoter at 5’end and tail oilgo sequence at 3’end with the following final sequences:

sgRNA3: TAATACGACTCACTATAGGgactgcccaatgccccgcaGTTTTAGAGCTAGAA

sgRNA4: TAATACGACTCACTATAGggcgagactgcttctcaaggGTTTTAGAGCTAGAA

NT gRNA: TAATACGACTCACTATAGggggaggcgttcggccacagGTTTTAGAGCTAGAA

sgRNAs were synthesized, as described in the studies by Medishetti et al and Sorlien.^18,19^ Briefly, templates for sgRNA3 and sgRNA4 were obtained by carrying out overlap PCR using T7 promoter forward primer and tail oligo reverse primer. The product was ethanol precipitated and in vitro transcribed using HiScribe™ T7 RNA synthesis kit (E2040S, New England Biolabs, Massachusetts, USA) to get the individual sgRNAs. A ccn2a guide RNA mix (sg3+sg4) or NT gRNA gRNA was allowed to form a complex with Cas 9 protein by incubating at 37°C for 10 minutes. This ribonucleoprotein (RNP) complex was microinjected (effective concentration of guides 1 ng per embryo along with 400 ng of Cas 9 protein) in one cell stage embryos, followed by incubation of the embryos at 28°C. The indels were visualized using the heteroduplex mobility assay (HMA) on 10% native PAGE after 24 hours post microinjection. Differential amplicon migration as compared to wild type control indicated presence of indels and confirmed effective editing in vivo.

### Phenotypic characterization of the *ccn2a* F0 knockout zebrafish larvae

Phenotypes were observed and imaged at 5dpf using bright-field microscopy (Zeiss Stereo Discovery.V8) using 3.5x magnification and as per published methodology.^20^

### Alcian blue staining

At 6.5dpf, zebrafish larvae were fixed in 4% paraformaldehyde (PFA) overnight. Subsequently, Alcian blue staining was performed as per the method described in earlier publication.^21^ Briefly, fixed embryos were washed three times with phosphate-buffered saline (PBS) for 10 minutes and the with 50% ethanol for 30 minutes. After ethanol wash, embryos were stained overnight at room temperature with 0.4% alcian blue prepared in 20mM MgCl_2_ and 70% ethanol. After staining, embryos were washed with 70% ethanol/10 mM MgCl_2_, 50% ethanol/10 mM MgCl_2_ and 25% ethanol, respectively. To reduce staining of soft tissues, embryos were bleached with 3% H_2_O_2_ and 2% KOH for 15 minutes. Embryos were washed with 25% glycerol and 0.25% KOH and stored in 50% glycerol in 0.25% KOH. Stained embryos were positioned in this storage solution in slits of the agarose mould and the head was photographed in a ventral–dorsal view. The defects in facial cartilage (Ceratobranchial pairs, Meckel’s cartilage, Ceratohyal) were visualized and images were compared with the wild type and NT controls.

### Quantitative real-time qPCR analysis

Total RNA was isolated using Trizol from 50 larvae at 5dpf for individual batch of injectants in n=5 experimental sets. cDNA was synthesized with 600 ng of total RNA using PrimeScript™ RT reagent Kit (RR037A-Takara Bio, Kusatsu, Japan) in a reaction volume of 10µl. qPCR was performed using TB Green mix on QuantStudio5 (Applied Biosystems, California, USA). Data were analyzed using Ct method (ΔΔCt) and normalized to *GAPDH* reference gene. 0.3µL of cDNA was used per reaction with zebra fish specific exonic primers for each of the genes studied that is *ccn2a, rac1a, rhoAa, col2a1a, sp7, runx2a and gapdh*. Relative gene expression was plotted for WT control, nontargeting (NT) injectant control and *ccn2a* guide injectants using Graph Pad Prism 5. The statistical significance was calculated using unpaired t-test.

## Results

### Clinical description

Family 1: We ascertained consanguineously marriedcouple with five pregnancies. Two living children (proband 1 and 2) have a kyphomelic dysplasia. They lost an affected female child and an affected pregnancy was medically terminated. They have a healthy daughter too. A boy and girl were affected in this family.

Family 2: A second consanguineous (third degree) family had four pregnancies that included proband 3 (P3) with a kyphomelic dysplasia. The couple had an earlier medical abortion of a similarly affected fetus and have two healthy children.

The clinical and radiographic findings are enumerated in table 1 and illustrated in figures 1-3.

**Table 1:** Summary of clinical and genetic findings in individuals with *CCN2* variants and kyphomelic dysplasia.

| Family ID | Family 1 |  | Family 2 |
| --- | --- | --- | --- |
| Subject ID | P1 | P2 | P3 |
| **General characteristics** |  |  |  |
| Gender | Male | Female | Female |
| Consanguinity | + | + | + |
| **Anthropometry** |  |  |  |
| Height in cm (SD) | 153 cm (-2.03) | 118cm (-3.6) | 76 cm (-5.5) at 3.5 years |
| Weight in kg (SD) | 41 kg (-1.91) | 25 kg (-2.18) | 14 kg (+0.06) at 3.5 years |
| Head circumference in cm (SD) | 52.5 cm (-1.61) | 50.5cm (-1.71) | 49 cm (-2.94) |
| **Clinical findings** |  |  |  |
| Short stature | + | + | + |
| Microcephaly | - | - | + |
| Bitemporal narrowing | + | + | + |
| Posteriorly placed ears | + | + | + |
| Deviated nasal septum | + | - | + |
| Micrognathia | + | + | + |
| Retrognathia | + | + | + |
| Microstomia | + | + | + |
| Cleft palate/ bifid uvula | + | + | + |
| Crowded teeth | + | + | + |
| Muscle wasting | + | + | + |
| **Skeletal findings** |  |  |  |
| Scoliosis | + | + | - |
| Platyspondyly | + | + | + |
| Short and broad pelvis | + | + | - |
| Coxa vara | + | + | + |
| Epiphyseal dysplasia | + | + | + |
| Metaphyseal dysplasia | + | + | + |
| Elbow joint dislocation | + | + | + |
| Bent radius and ulna | + | + | + |
| Limited elbow extension/joint mobility | - | + | + |
| Femoral bending (kyphomelia) | + | + | + |
| Bowing of tibia and fibula | + | - | + |
| Mobile patella | + | - | + |
| Windswept deformity | + | - | - |
| Genu valgum | - | + | + |
| Limited knee flexion | + | + | + |
| Bilateral pes planus | + | - | - |
| Pes cavus | - | - | + |
| Broad great toe | + | + | + |
| **Molecular findings in *CCN2*** |  |  |  |
| Nucleotide change (NM_001901.4) | c.443G>A | c.443G>A | c.779_786del |
| Amino acid change (NP_001892.2) | p.(Cys148Tyr) | p.(Cys148Tyr) | p.(Pro260LeufsTer7) |
| Location | Exon 3 | Exon 3 | Exon 5 |
| Zygosity | Homozygous | Homozygous | Homozygous |
| Parental status | Heterozygous | Heterozygous | Heterozygous |
| Variant effect | Missense | Missense | Frameshift |
| +: Present; -: Absent |  |  |  |

**Figure 1:**
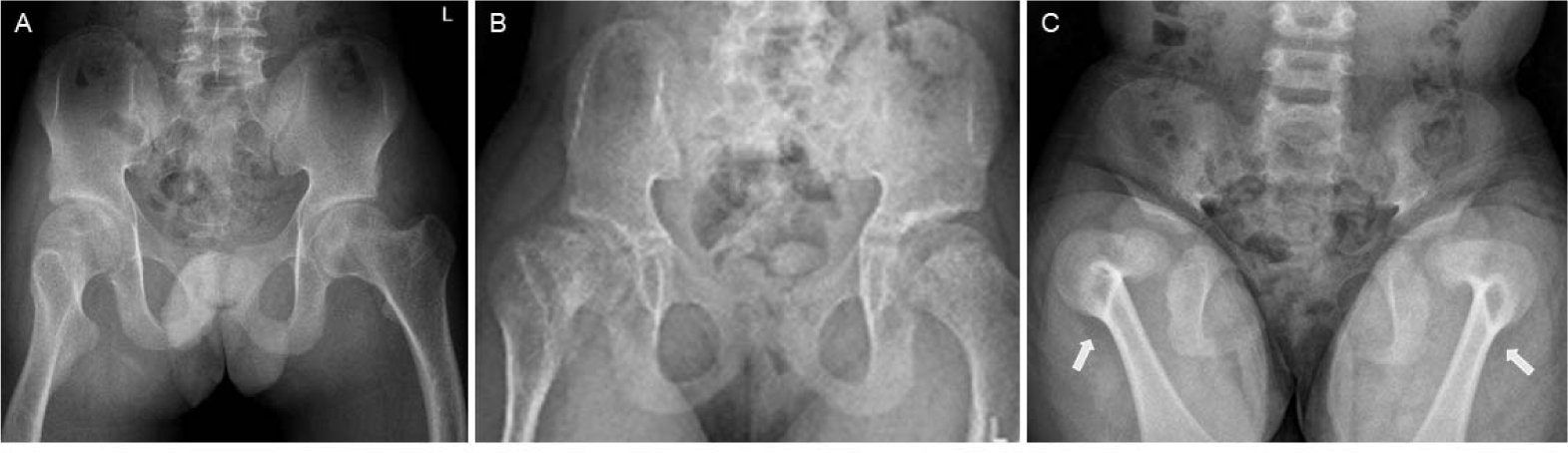
Kyphomelic femora in the participants. Radiographs of pelvis [proband 1 (A), proband 2 (B) and proband (C)] show short and broad pelvis, coxa vara and reduced hip joint space in all. Kyphotic femora are seen in all of them but is more prominent (arrows) in the younger proband 3 (C).

**Figure 2:**
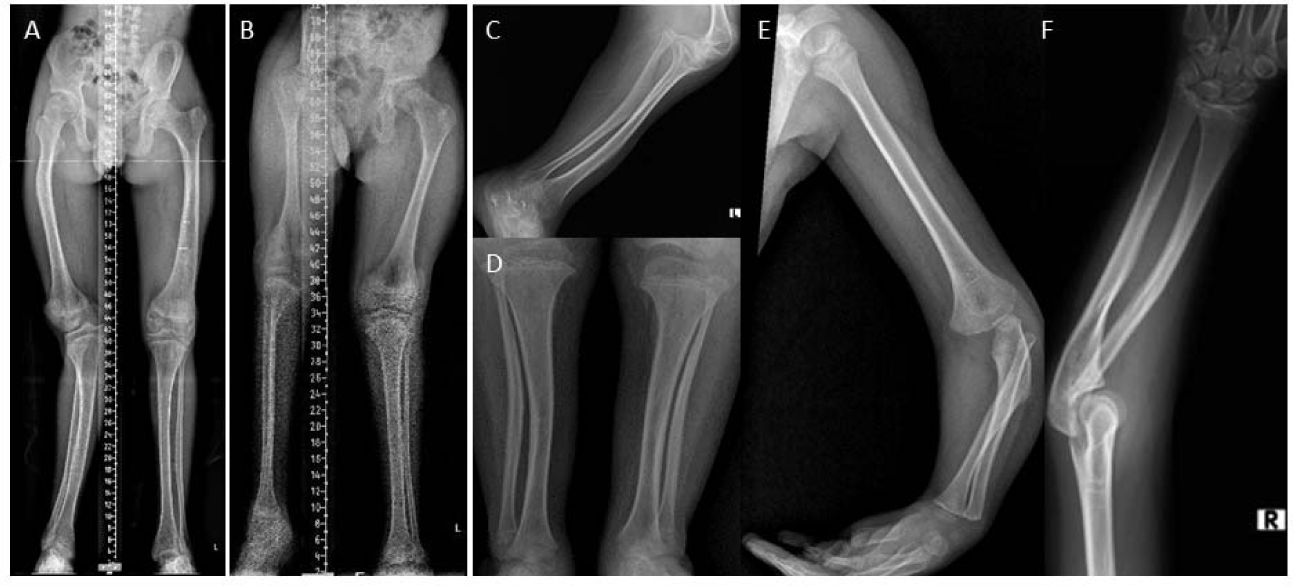
Bowing of long bones in the participants. Radiographs of limbs for probands: proband 1 (A), proband 2 (B) and proband 3 (C, D, E and F) show bowing of long bones. Bowing of femur (A and B), bowing of the tibia and fibula (A, C, D) can be observed. Irregular epiphyses and metaphyses along with flaring of metaphyses at the knee joint can be noted (D).

**Figure 3:**
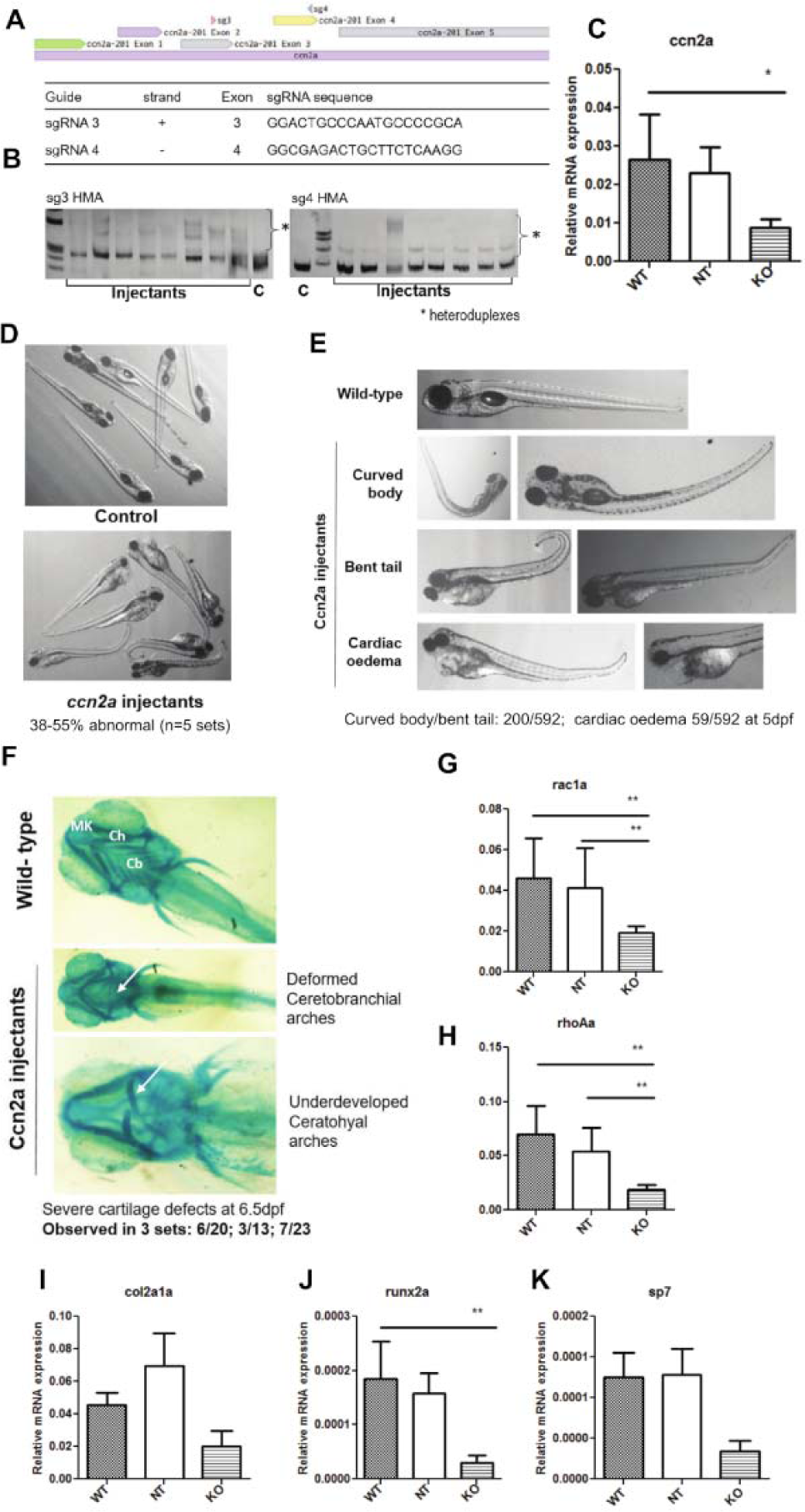
Phenotypes in the ccn2a knockout zebrafish. (A) The genomic locus of the ccn2a gene in the zebrafish genome with the location of the selected sgRNA target sites indicated. The sequences of the two sgRNAs used in the study are provided in the table. (B) Assessment of editing efficiency for each sgRNA at 24 hours post injection, by a heteroduplex mobility assay (HMA). (C) Relative expression levels of the ccn2a mRNA in the controls and injectants (crispants). (D) Bright field image of a group of ccn2a crispants and matched control injectants from the same experiment to illustrate the extent of physical phenotypes. (E) Representative images of observed phenotypes as labeled, among the ccn2a crispants. (F) Alcian blue stained images of wildtype and representative examples of ccn2a crispants, showing cartilage deformities. Cartilage elements labeled: MK: Meckel’s cartilage; Ch: ceratohyal; Cb: ceratobranchial pairs. G-K. Relative mRNA levels of candidate genes including ccn2a, col2a1a (chondrocyte marker), rac1a and rho1a (palatogenesis markers) and sp7 (osteoblast marker), in the ccn2a crispants as compared to the controls. Results from 5 independent experiments are quantified.

### Molecular findings

Exome sequencing in three probands from the two unrelated families were non-diagnostic. In Family 1, a shared biallelic non-synonymous missense variant, c.443G>A; p.(Cys148Tyr), located in exon 3 of the *CCN2* (NM_001901.4; NP_001892.2), was identified through duo exome sequencing in both siblings. This variant is absent in gnomAD (V3.1.2) and our in-house data of 3188 exomes. It is predicted to substitute a cysteine residue with tyrosine in the von Willebrand factor type C (VWC) domain of the CCN2 protein. Multiple sequence analysis performed using the Clustal Omega tool revealed conservation of the cysteine amino acid residue across several vertebrate species (Figure 4). In silico pathogenicity prediction tools, including CADD phred: 31.00 and REVEL: 0.967, predicted that the variant is disease-causing. Additionally, the AlphaMissense score for the variant p.(Cys148Tyr) is 0.996, further indicating its potential pathogenicity. Variant validation and segregation analysis using Sanger sequencing confirmed that the variants are present in heterozygous state in the parents and found absent in the unaffected sibling.

**Figure 4:**
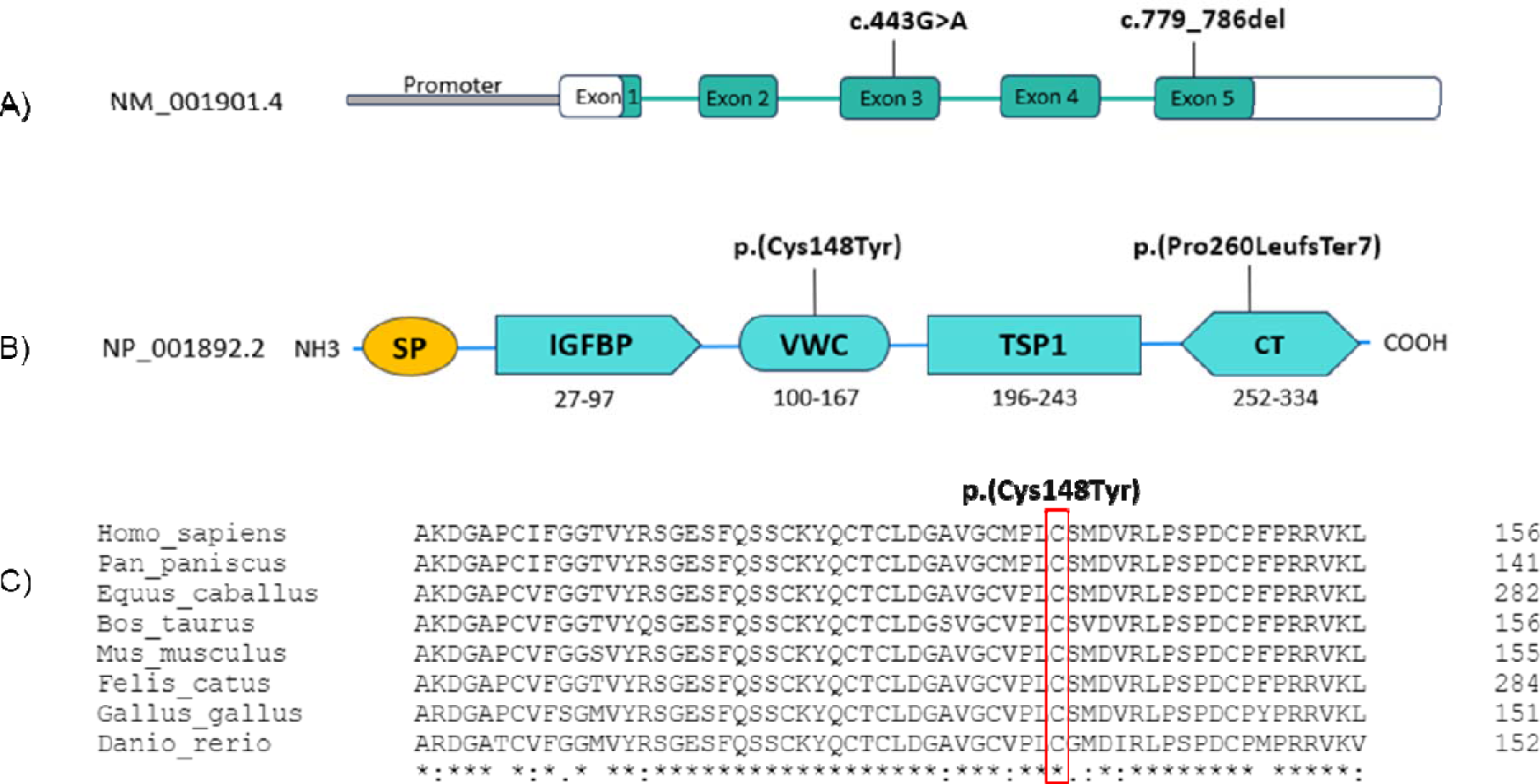
(A) Schematic illustration of structure of CCN2 gene with biallelic variants identified in the study (B) Structure of CCN2 proteins with four domains (IGFBP, VWC, TSP-1 and CT) and variants detected in the study. The Arabic numerals display the size of each domain. SP, signal peptide; IGFBP, insulin-like growth factor-binding protein; VWC, Von Willebrand factor; TSP-1, thrombospondin type-1; CT, cysteine knot. (C) Multiple sequence alignment of p.(Cys148Tyr) in CCN2 gene indicates that cysteine residue at 148 position is conserved among different vertebrates. An asterisk (*) represents fully conserved residue, colon (:) represents residues with strongly similar properties, and period (.) represents residues with weakly similar properties.

In family 2, we identified a novel homozygous frameshift variant, c.779_786del; p.(Pro260LeufsTer7), located in exon 5 of *CCN2*. Exon 5 of *CCN2* encodes for carboxyterminal domain, which is critical for interaction with cell surface integrins. ^22^ This variant is likely to result in either the formation of a truncated protein or the transcript may undergo nonsense mediated mRNA decay which might perturb the proper functioning of the protein. Sanger sequencing confirmed the heterozygous status of the parents. Detailed clinical, radiographic, and molecular findings of probands are summarized in table 1.

### Functional studies on zebrafish

The zebrafish genome contains two *CCN2* orthologs *ccn2a* and *ccn2b*, among which we chose to work with *ccn2a* for two primary reasons: a) *ccn2a* is phylogenetically closer to human *CCN2*, and b) *ccn2b* is not well expressed during early development (Figure 5).^23^ In order to determine whether the loss of *ccn2a* function leads to impaired skeletal development, we created F0 knockouts of *ccn2a* in zebrafish as reported previously.^24^ We designed four guides targeting the *ccn2a* locus, spanning the entire coding sequence and tested these for efficacy of editing in vivo, by injecting the Cas9-sgRNA RNPs into 1-cell stage embryos and performing genotyping at 24hpi by HMA PCR. Two of the four guides were chosen based on efficacy and used at 1ng guide RNA mix per embryo for all subsequent experiments (Figure 3A and B). Two controls were used for each experiment, uninjected embryos (WT) and injections with a not targeting guide RNA (NT) injected at the same amount as the ccn2a guides. A significant decrease in *ccn2a* mRNA was observed in the crispants as compared to the NT and WT controls, thus confirming *ccn2a* editing (Figure 3C). *ccn2a* crispants showed significant abnormalities such as body curvature and bent tail which are characteristic of defects in early skeletal development (Figure 3D). A small but significant fraction of crispants showed severe cardiac edema. The number of crispants showing these phenotypes was quantified in each experiment, and representative images are shown (Figure 3E). The ccn2a crispants showed significant defects in cartilage formation in the craniofacial region as seen by Alcian blue staining at 6.5dpf (Figure 3F). They had underdeveloped developed ceratohyal arches, bent ceratobranchial arches and misshapen Meckel cartilage.

**Figure 5:**
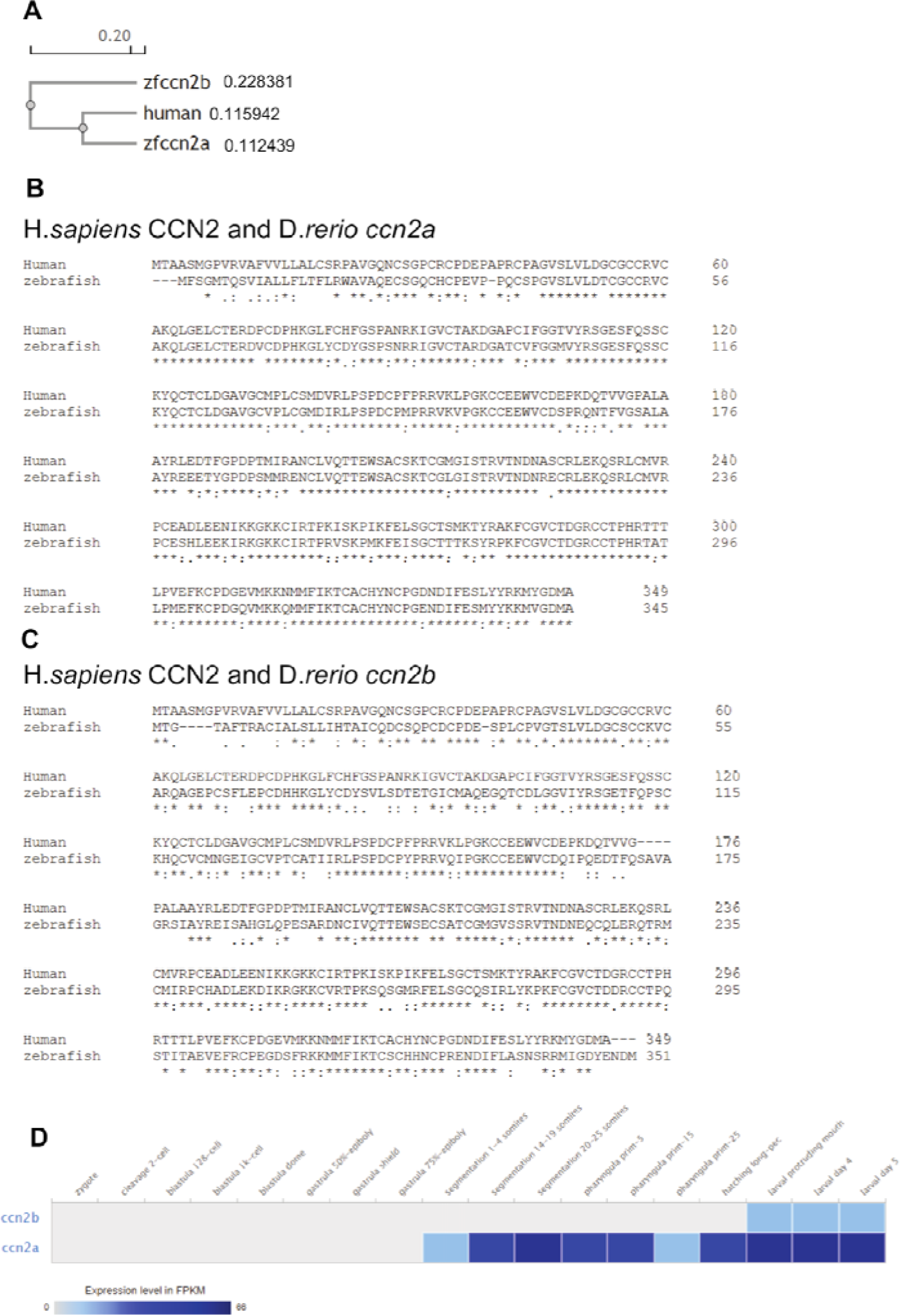
Phylogram showing the evolutionary relationship (A) and protein sequence alignments (B and C), between the human CCN2 and zebrafish Ccn2a and Ccn2b *protein* sequences. D. Expression data of zebrafish *ccn2a* and *ccn2b* mRNA from the transcriptional profiling of zebrafish developmental stages.

To further confirm a role for *ccn2a* in early cartilage and bone formation, we have examined the levels of established skeletal marker genes such as *col2a1a* (chondrocyte marker), *rac1a* and *rhoAa* (palatogenesis markers), *sp7* and *runx2a* (osteoblast markers). The *ccn2a* crispants showed a significant decline in the levels of all of these markers, as is expected based on the severe phenotypic defects (Figures 3G-K).

The crispants also showed poor survival beyond 7dpf and few survived to adulthood. These adult crispants (F0 KO) showed defects in mineralization and bone structure in specific locations with known endochondral ossification (Figure 6C) such as missing structures in the tail region (hypural bones) and abnormal trunk curvature (Figure 6A-B). The level of knockdown of *ccn2a* in these adults was confirmed by measuring the mRNA level from trunk tissue and was found to be significantly reduced (Figure 6C).

**Figure 6:**
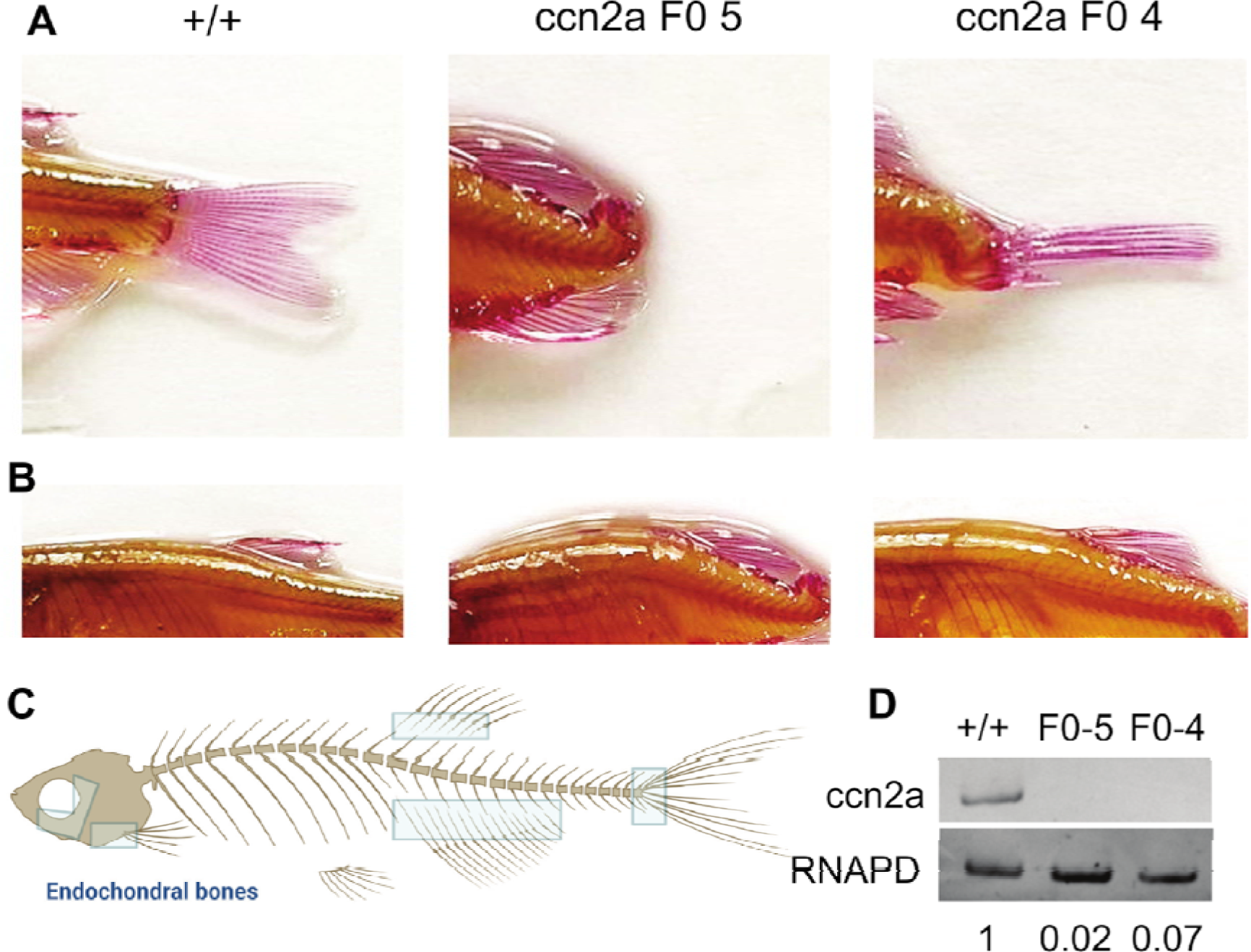
Phenotypes in the *ccn2a* crispant (F0) knockout adult zebrafish. Images of the tail section (A) and trunk (B) of adult zebrafish, fixed, bleached and stained with Alizarin Red to visualize bone mineralization. (C) Schematic of the zebrafish skeleton with rectangles highlighting regions with endochondral bones. (D) Levels of *ccn2a* mRNA measured in tissue isolated from the trunk of the fish in (A) by semi-quantitative RT-PCR.

## Discussion

We describe an autosomal recessive kyphomelic dysplasia in multiple affected individuals from two unrelated consanguineous families. We identified two homozygous variants (missense and frameshift) in *CCN2*, which encodes a protein involved in proliferation and differentiation of chondrocytes. Further, we investigated the consequences of loss of function of *CCN2* in zebrafish models. Mice and zebrafish models replicate the human phenotype.

Kyphomelic dysplasia is a heterogeneous disorder, with the most notable feature being the presence of bowing of long bones, particularly severely angulated femurs.^1^ Kyphomelic femur is usually prominent in children but becomes less pronounced progressively.^1,2,7^ Though six offsprings were affected in the two families described here, we could document kyphomelic femora in all three affected children who were available for clinical evaluation. Our observations are also in concordance with the literature as the proband 3 showed more acute angulation at 3.5 years than the older probands in family 1. Additionally, observation of femoral bending in the antenatal scans in several affected members (including medical abortions) in this family supports the age-related reduction in severity of kyphomelia in this condition. We observed facial dysmorphism in all affected children. These include microstomia, micro-retrognathia, posteriorly placed ears and bitemporal narrowing. These features are variably noted in the historical reports of kyphomelic dysplasia, though genetic defects were not reported in them^1,6–8^.

A relatively milder phenotype in the family 1 may partially be attributed to the type of variant in *CCN2*. Though it is too early to speculate in view of only two families, the missense variant might have contributed to the less severe kyphomelia and lesser reduction in height in the family 1. It is worth noting that the family 2 had a predicted truncating variant (in the last exon which might escape nonsense mediated decay of mRNA) and had more acute femoral bending and severely reduced height. As we are unable to study the CCN2 protein and its function in patient cell lines, this remains a hypothesis that needs to be tested in more patients.

Kyphomelic dysplasia can frequently be misdiagnosed as campomelic dysplasia, a well-known condition that also presents with milder bowing of the femur. However, consistent and distinguishing findings in campomelic dysplasia are severe tibial affliction, the small/absent scapula, and hypoplastic pedicles of the thoracic vertebrae.^25^ Additionally, campomelic dysplasia is usually due to heterozygous disease-causing variants in the *SOX9* gene.^26^ In view of the more severely bent femora, we would like to use the term kyphomelia in description of the femora here, while we agree the boundaries are indistinct between kyphomelia and campomelia.

We considered other conditions that may be mistaken for kyphomelic dysplasia: Stuve–Wiedemann syndrome, Schwartz-Jampel syndrome type 1, or metaphyseal chondrodysplasia, McKusick type. These disorders share bowing of the lower limbs. However, they result from biallelic variants in the *LIFR*, *HSPG2*, and *RMRP* genes, respectively and our genomic test results did not support these diagnoses.^6,7,27^

The genetic etiology of kyphomelic dysplasia has remained poorly understood as yet. Although numerous reports have described about 23 patients with kyphomelic dysplasia, none of these have provided molecular etiology of the disorder.^6,28–30^ It is possible that some of these might represent other genetic disorders with bent long bones and even have a non-genetic etiology. Nevertheless, kyphomelic dysplasia is typically regarded as an autosomal recessive condition in the reported cases.^2,8^ In a recent study Itai et al., described *de novo* heterozygous variants in *KIF5B* can lead to kyphomelic dysplasia in four individuals inherited in an autosomal dominant manner.^4^ Affected individuals had short stature, bowing of limbs and facial dysmorphism including bitemporal constriction, arched eyebrow, hypertelorism, proptosis, ptosis, midface hypoplasia, micrognathia and cleft palate. However, our present study, through pedigree analysis and segregation of *CCN2* variants confirms autosomal recessive inheritance of this disorder.

CCN2/CTGF is a multifunctional protein spanning 349 amino acids, belonging to cysteine rich CCN protein family, which show conservation of all cysteine residues.^31^ It comprises of four modules, namely: IGF (insulin like growth factor)-binding protein-like (IGFBP), von Willebrand factor type C (VWC), thrombospondin type 1 repeat (TSP1) and C-terminal cystine knot (CT).^32^ Each of these modules serve distinct functions. *CCN2 gene* has five exons. This study identified a missense variant (c.443G>A); p.(Cys148Tyr) and a frameshift variant (c.779_786del); p.(Pro260LeufsTer7) which reside in exon 3 and exon 5 of *CCN2* which encode for VWC and CT domains of the protein respectively (Figure 4). The module 4 of *CCN2*, VWC interacts with signalling molecules such as transforming growth factorβ (TGFβ), bone morphogenetic protein (BMP) and Wnt. During chondrogenesis, coordinated involvement of these signalling molecules or growth factors are required for proper development.^33,34^ A missense substitution of evolutionary conserved cysteine residue in VWC domain, may lead to loss of protein function. The C-terminal module participates in binding to specific integrins. This interaction with cell surface integrins contributes to cell adhesion, migration, and deposition of extracellular matrix proteins.^22^ A homozygous frameshift variant in the CT domain may result in the transcript undergoing nonsense-mediated decay or forming a non-functional truncated protein, potentially impairing protein function. Furthermore, both variants are not present in the gnomAD database or our in-house cohort of 3188 exome data. Additionally, *in silico* tools predicting the detected variants to be disease-causing.

Several in vitro studies have confirmed that CCN2 acts as a growth factor, promoting the proliferation, differentiation, and maturation of chondrocytes and osteoblasts.^10,11,35^ In vivo studies using mice models, demonstrated that *Ccn2* deficient mice manifest skeletal developmental abnormality that comprises craniofacial abnormalities such as shortened mandibles resulting from deformation in Meckel’s cartilage, domed cranial vault and distorted ethmoid bones along with secondary cleft palate. Additionally, bends and kinks were apparent in the long bones (radius, ulna, tibia and fibula). *Ccn2* deficient mice also exhibited perinatal lethality due to respiratory failure primarily attributed to short and bent sterna and kinked ribs. These findings closely mirrored the clinical manifestations observed in our study.^9,12,36–38^ In studies in knockout mice, absence of CCN2 is also reported to inhibit palatal shelf elevation from the vertical to horizontal position thus demonstrating its importance in mammalian palatogenesis. This can be observed as the occurrence of cleft palate in humans, which is evident in all probands in the study.^39^ Mice homozygous for deletion of *Ctgf* gene die soon after birth.

Zebrafish have increasingly emerged as efficient and economical models for genetic research, leveraging the functionality of the CRISPR/Cas9 system. Given the high conservation of fundamental signalling pathways and cellular processes involved in skeletal development from fish to humans, zebrafish serve as valuable models for studying skeletal disorders.^36,40^ Numerous human skeletal disorders have been successfully replicated in zebrafish models.^41^ Using *ccn2a* F0 knock out crispant in zebrafish, we demonstrate that deficiency of CCN2 can result in skeletal manifestations.

In zebrafish *ccn2a* has been shown to play important role in heart regeneration after cardiac injury.^42^ It has also been reported to have regenerative activity and is required for spinal cord regeneration, pro-regenerative activity of *ccn2a* maps to its C-terminal domains,^42,43^ however its function in early skeletal development in zebrafish has not yet been studied. The previous reports using the ccn2a-/- line did not report any major developmental or skeletal phenotypes.^44^ We believe this could be due to genetic compensation mechanisms which may arise due to the severe defects associated with loss of *ccn2a*, as has been reported previously for select genes,^45^ especially given that significant compensatory upregulation of *ccn2b* was observed in the *ccn2a* -/- line. To circumvent these issues, we have therefore used an effective F0 knockout strategy described by Wu et al 2018, to create a loss of *ccn2a* function and study the impact during larval stages on cartilage formation and subsequently mineralization.^24^ This method has been reported to recapitulate knockout phenotypes in more than 90% of F0 embryos (crispants), with persistence well into adulthood.

In *ccn2a* crispants, we observed skeletal developmental abnormalities similar to the phenotypes observed in patients with *CCN2* variants which led to skeletal dysplasia and cleft palate. A significant decrease in osteogenic markers like *Col2a1*, *sp7*, and *runx2* was noted during zebrafish development. This decline may be attributed to reduced CCN2 expression, which plays a pivotal role in inducing these markers through various pathways.^38^ The comparison of phenotypes observed among zebrafish, mice and humans are described in table 2.

**Table 2:** Comparison of phenotypes observed in zebrafish, mice and human due to loss of function of *CCN2* gene.

| Phenotypes observed due to loss of function of *CCN2* | *ccn2a* knockout zebrafish | *Ccn2* deficient mice | Affected individuals |
| --- | --- | --- | --- |
| Craniofacial abnormalities | Underdeveloped ceratohyal arches, bent ceratobranchial arches and misshapen Meckel's cartilage | Shortened mandibles, deformed Meckel's cartilage and ethmoid bones, domed cranial vault, and secondary cleft palate | Bitemporal narrowing, posteriorly placed ears, deviated nasal septum, short/bifid uvula, crowded teeth, micrognathia, microstomia and retrognathia |
| Vertebral/ ribs abnormalities | Altered body curvature | Short and bent sterna and kinked ribs | Platyspondyly, mild scoliosis and irregularities at vertebral ends |
| Upper limb deformities | - | Bends in radius, ulna | Radial head dislocation and bent radius and ulna |
| Lower limb deformities | Bent or missing tail | Curved tibia and fibula | Kyphomelic femur, bent tibia and fibula, broad great toes, genu valgum and windswept deformity |
| Epi-metaphyseal dysplasia | - | - | Irregular epiphyses and irregular and flared metaphyses |
| References | Present study | Ivkovic et al., 2003 | Present study |
| - : Absent/not known |  |  |  |

We acknowledge certain limitations in our study. We are unable to provide cellular effects of CCN2 variants in these patients, such as immunocytofluorescence and western blot analyses. These analyses would have allowed us to examine expression patterns of CCN2 variants and osteogenic markers compared to control samples. We report only on F0 knockouts in this study, as creating zebrafish lines expressing mutant Ccn2a proteins would be technically challenging, time-consuming and financially demanding, and beyond the scope of our current expertise. Unavailability of a resolved CCN2 protein structure hindered the protein modelling for assessing interactions affected due to substitution of cysteine with tyrosine residue at position 148.

In summary, we present two unrelated families with multiple affected individuals with an autosomal recessive kyphomelic dysplasia, resulting from likely loss of function of the *CCN2*. The mice knockouts have already been described to have bone dysplasia akin to the human phenotype described here. We also show zebrafish knockouts for *ccn2* recapitulate human skeletal dysplasia. However, investigation of additional patients and cellular studies are necessary to establish the gene-disease relationship.

## Data availability

The data that support the findings of this study are available from the corresponding author upon reasonable request.

## Acknowledgement

We thank the patients and their families for participating in the study. We would like to acknowledge the invaluable contributions of Dr Alka Ekbote to this manuscript. She played a significant role in conceptualization, formal analysis, and investigation. Her contributions have enriched the outcome of this work. Although Dr Alka Ekbote is no longer with us, her legacy continues to inspire our team.

## Author contribution

Conceptualization: S.S., H.S., S.D., A.E., A.S., G.S.B., K.M.G.; Data curation: S.S., G.S.B., H.S., A.E., A.S., K.M.G.; Formal analysis: S.D., V.M., A.E., G.S.B., N.S., N.Z.P., R.M., A.S.; Funding acquisition: S.D., A.S., K.M.G; Investigation: S.S., A.E., S.D., V.M., N.S., N.Z.P., R.M., M.T.A., H.S., A.S., G.S.B.; Methodology: A.S., N.S., S.D., A.E., N.Z.P., R.M., G.S.B., K.M.G.; Project Administration: K.M.G.; Resources: S.D., V.M., A.E., A.S., K.M.G., Supervision: S.D., A.S., G.S.B., K.M.G.; Writing the first draft: S.S., A.E., S.D., A.S., K.M.G.; Writing-review & editing: S.S., S.D., N.S., H.S., V.M., M.T.A., N.Z.P., R.M., G.S.B., A.S., K.M.G. All authors have edited the manuscript drafts and revisions and approved the final version of the manuscript.

## Ethical declaration

Informed consents were obtained from the participants for the study and publication of clinical photographs. The research protocol is approved by the Institutional Ethics Committee, Kasturba Medical College and Hospital, Manipal (IEC: 363/2020).

## Web resources

PRIMER 3 v.4.1.0, http://primer3.ut.ee/

Ensembl, https://asia.ensembl.org/index.html

Mutation Taster, http://www.mutationtaster.org/

CADD Phred, https://cadd.gs.washington.edu/

MCAP, http://bejerano.stanford.edu/mcap/

REVEL, https://genome.ucsc.edu/cgi-bin/hgTrackUi?db=hg19&g=revel

SIFT Indel, https://sift.bii.a-star.edu.sg/www/SIFT_indels2.html

Clustal Omega, https://www.ebi.ac.uk/jdispatcher/msa/clustalo

AlphaMissense: https://alphamissense.hegelab.org/search

Online Mendelian Inheritance in Man (OMIM): https://www.omim.org/

gnomAD, https://gnomad.broadinstitute.org/

HPO, https://hpo.jax.org/app/

LOVD, https://www.lovd.nl/

ANNOVAR, http://annovar.openbioinformatics.org/

